# Previous SARS-CoV-2 infection or a third dose of vaccine elicited cross-variant neutralizing antibodies in vaccinated solid organ transplant recipients

**DOI:** 10.1101/2022.04.13.22273829

**Authors:** Chih-Chao Chang, George Vlad, Elena Rodica Vasilescu, Ping Li, Syed A. Husain, Elaine A. Silvia, David J Cohen, Lloyd E. Ratner, Wei-Zen Sun, Sumit Mohan, Nicole Suciu-Foca

**Affiliations:** Department of Pathology and Cell Biology, Columbia University Irving Medical Center, New York, USA; Department of Medicine, Division of Nephrology, Columbia University Irving Medical Center, New York, USA; The Columbia University Renal Epidemiology (CURE) Group, New York, USA; Department of Surgery, Columbia University Irving Medical Center, New York, USA; Department of Anesthesiology, National Taiwan University Hospital, Taipei, Taiwan

## Abstract

The SARS-CoV-2 pandemic poses a great threat to global health, particularly in solid organ transplant recipients (SOTRs). Although a 3-dose mRNA vaccination protocol has been implemented for the majority of SOTRs, its effectiveness was still largely unknown. We analyzed 113 vaccinated SOTRs, and 30 healthy controls (HCs), some of whom had recovered from COVID, for their immune responses against the original vaccine strain and variants of concern (VOC), including the highly mutated-omicron variant. Here, we report that 3 doses of the mRNA vaccine had only a modest effect in eliciting anti-viral responses against all viral strains in the fully vaccinated SOTRs who did not contract the virus. Only 34.0% (16/47) of this group of patients demonstrated both detectable anti-RBD IgG and neutralization activities against alpha, beta, and delta variants, and only 8.5% (4/47) of them showed additional omicron-neutralizing capacities. In contrast, 79.5% (35/44) of the vaccinated recovered-SOTRs demonstrated both higher anti-RBD IgG levels and neutralizing activities against all VOC, including omicron. These findings illustrate a significant impact of previous infection on the development of anti-COVID immune responses in vaccinated SOTRs and highlight the need for alternative strategies to protect a subset of a lesser-vaccine responsive population.

## 1.1 Introduction

Solid organ transplant recipients (SOTRs) are at risk for severe COVID-19, because of the use of immunosuppression and/or impaired immune defenses caused by underlying diseases. Patients with SARS-CoV-2 are thought to have increased rates of bacterial and fungal superinfections^1^. The situation is further complicated by the fact that severe SARS-CoV-2 infection are associated with cytokine storms, an event marked by uncontrolled release of inflammatory cytokines in infected patients^2^. Disease severity and mortality among SOTRs with SAR-CoV-2, were found to be quite high in the early period of pandemic^3,4^, but have gradually trended down, as greater access to testing and better therapeutic intervention have been implemented^5^.

Although the use of mRNA COVID vaccines provides relief for the general population by preventing severity of disease and/or contraction of the virus, the effectiveness of the standard 2-dose vaccination has been found to be insufficient for SOTRs^6,7^. In particular, the emergence of many SARS-CoV-2 variants, including variants of concern (VOC), led to the authorization of using an additional 3^rd^ dose of the vaccine as part of the primary immunization series. Recent studies^8-11^ indicate that the 3^rd^ dose could boost the levels of anti-SARS-CoV-2 antibodies, although neutralizing activities against the original vaccine strain and some VOC were significantly weaker in SOTRs when compared to healthy controls^11^.

Almost 80 million people in United States, including many SOTRs, contracted SARS-CoV-2 since March 2020. Like the general U.S population, the majority of SOTRs who had recovered from SARS-CoV-2 received at least two doses of the vaccine. It is abundantly clear that vaccination increases both the levels of anti-SARS-CoV-2 antibodies and cross-variant neutralizing capacities in individuals, regardless of the status of SARS-CoV-2 infection^12-14^.

We previously reported that a majority of kidney transplant patients who contracted SARS-CoV-2 had retained anti-RBD IgG, presumably the protective antibodies, but lost anti-nucleocapsid IgG antibodies after a prolonged period of time (200 days)^15^. In the current study, we investigate if vaccinated SOTRs, who either recovered from COVID-19 (referred to as recovered-SOTRs) or had not contracted the virus (referred to as COVID-naïve), were capable of mounting efficient humoral responses against the vaccine strain or VOC, including the current highly mutated-omicron variant.

## Materials and Methods

### 2.1 Human Subjects, Patients Privacy Protection

This retrospective study was approved by a protocol (AAAT3602) from the Institutional Review Board at Columbia University Irving Medical Center (CUIMC). In all, a total of 113 solid organ transplant recipients (SOTRs), who returned to CUIMC for patient care from March 2020 to January 2022 and had been vaccinated with 2-3 doses of SARS-CoV2 mRNA vaccines (either Moderna mRNA-1273 or Pfizer-BNT 162b2) were identified through our medical record system. COVID-19 diagnosis was confirmed by a positive SARS-CoV-2’s RT-PCR test on nasal swab samples. One group selected (recovered SOTR, n = 44) were patients who recovered from a previous COVID infection. The other group, (COVID-naïve SOTR, n = 69), were those who had not been infected, and were confirmed as seronegative for anti-nucleocapsid IgG antibodies. Also included were 30 healthy controls (HCs), either healthcare workers or volunteers. Verbal informed consent was given by HCs.

### 2.2 Laboratory finding and sample collection

Authorization for use of de-identified SOTRs specimens from clinical care that would otherwise be discarded, and use of de-identified laboratory results of SOTRs were approved by a waiver (AAAP2200) through a HIPPA Guideline. Serum samples of SOTRs, previously collected for patient care were identified by *Histotrac* Software from the Serum Bank of the Immunogenetics Laboratory of CUIMC. Sera from HCs were prepared from whole blood via centrifugation, aliquoted, and frozen at -20°C.

### 2.3 Plasmids, cell lines, transfections and SARS-CoV-2 pseudoviral particles production

The following plasmids were obtained from Addgene Inc: pcDNA3.3_CoV2_B.1.1.7 (alpha variant, #170451), pcDNA3.3_CoV2_501V2 (1.351, beta variant, #170449), pcDNA3.3-SARS2-B.1.617.2 (delta variant, #172320) and pTwist-SARS-CoV-2 Δ18 B.1.1.529 (Omicron variant, #179907). All these plasmids encode spike proteins with c-terminal 18 aa deletion. The following plasmids were obtained from Bei Resources: pHDM-SARS-CoV-2-SpikeD614G (NR-53765), Lenti-pseudoviral packing plasmids, pHAGE-CMV-Luc2-IRES-ZsGreen-W (NR-52516), pHDM-Hgpm2 (NR-52517), pHDM-tatt11b (NR-52518) and pRC-CMV-rev1b (NR-52519). The human ACE2 stably transfected cell line, 293T-ACE2, was also from Bei Resources.

SARS-CoV-2’s pseudoviral particles were prepared from transfection of 293T cells with lentiviral packing plasmids, together with various SARS-CoV-2 Spike coding plasmids, according to a method described by Crawford^16^ et al. Plasmid DNA was purified by Qiagen EndoFree® plasmid kit; transfections were performed with lipofectamine® 3000 (Fisher Scientific, Pittsburgh, PA USA). All SARS-CoV-2 pseudotyped lentiviral particles were filtered through a 0.45 µm filter, concentrated by centrifugation through a 10% sucrose cushion, aliquoted, and stored at -80°C. The schematic structures of plasmids encoding for spike proteins of SARS-CoV-2’s pseudoviruses are shown in Figure S1.

### 2.4 SARS-CoV-2 pseudotyped neutralization assays

Neutralization assays were performed by incubating SARS-CoV-2 pseudovirus with serial dilutions (1:10 to 1:10,000) of sera and scored by the reduction in luciferase gene expression. In brief, 25 µl pseudovirus (3-5×10^5^ RLU) was incubated with an equal volume of diluted sera in a 96 well-plate at 37°C for 45 min; 40 µl of serum/virus mix were then transferred to wells of a cell culture 96 well-plate. Each well was pre-seeded with 1.5×10^4^ 293T-ACE2 cells in 60 µl medium (DMEM supplemented with 10% FCS) in the presence of 8 µg/ml polybrene. After 60 h post-infection, cells were collected for luciferase assays. The Promega Bright-Glo® Luciferase Assay System with Promega GloMax® Plate Reader was used for detection of luciferase activity. The qualitative analysis (% neutralization) is defined as 100*(1-(sample’s RLU - background RLU)/ (positive control’s RLU - negative control’s RLU)). IC50 (half maximum inhibitory concentration) values were calculated using nonlinear regression in GraphPad Prism 9.2.0.

### 2.5 Multiplexed magnetic bead-based assay for detection IgG antibodies against SARS-CoV-2’s viral antigens

The xMAP^®^ SARS-CoV-2 Multi-Antigen IgG system for simultaneous detection of viral targets (RBD, S1 and nucleocapsid viral proteins) was previously described^15^. The positivity of anti-viral IgG antibodies was pre-set by manufacturer as equal to or above 700 MFI.

### 2.6 Multiplexed magnetic bead-based assay for detection of neutralizing antibodies against the vaccine strain or VOC

Bio-Rad’s Bio-Plex Pro® Human SARS-CoV-2 Neutralization Assay was used for detection of neutralization activities with some modifications. In brief, multiplexed magnetic beads were prepared by mixing SARS-CoV-2 neutralization antibody 2-Plex (the original Spike 1 and RBD) with viral antigen coupled beads of Alpha S1, Beta S1, Delta RBD and Delta Spike Trimer (all from Bio-Rad). After the second wash, magnetic beads were mixed with 25 µl of serially diluted (1:5-1:1500) subject’s sera and incubated at room temperature (RT) for 30 min. with shaking. Biotin-labeled human ACE2 (25 µl) was then added to the reaction wells and incubated at RT for another 30 min. After 3 washes, ACE2 binding magnetic beads were incubated with 50 µl detection reagent (streptavidin-PE) at RT for 10 min with shaking. Re-suspended beads were transferred to a V-bottom 96 well-plate and run on a Luminex^®^200 Platform. Data (mean fluorescence intensity, MFI) were acquired using xPONENT^®^ Software and analyzed with Microsoft’s Excel Software. Manufacturer’s cutoff for positive neutralization was the percentage of inhibition above 10% with sera diluted at 1:5. Based on the results from Figure S2, our positive neutralization cutoff was set to equal or above 8% inhibition with sera diluted at 1:200 fold, or equal or above 30% inhibition with sera diluted at 1:20 fold.

### 2.7 Statistics

Statistical analyses and generation of the graphs were carried out using GraphPad Prism 9.0. Unpaired t test with Welch’s correction was used to compare two groups of variables. The Spearman correlation coefficient r was calculated for quantifying the association between continuous variables. Two-tailed *p* values were reported with *p* < 0.05 considered significant.

## Results

### 3.1 Description of the Study population

As shown in Table I, both COVID-naïve SOTRs and recovered-SOTRs shared similar demographics and clinical characteristics. They were similar in age (56.1 vs 61.0 yrs.), and they were mostly kidney transplant recipients (77.3.% vs 73.9%) who received organs mainly from deceased donors In addition, a majority of the recipients were tested within 3 years post-transplantation and received similar regimens (prednisone, calcineurin inhibitors, mTOR inhibitors and anti-metabolites) for maintenance immunosuppression. Post-transplant monitoring of organ recipients indicated that a vast majority (>90%) had no donor specific anti-HLA antibodies in their sera at the time of testing, suggesting that transplanted grafts were stable at the time of vaccination and sampling. A majority of recovered-SOTRs (33/44) as well as recovered-HCs (6/8) contracted SARS-CoV-2 during the first wave of the pandemic as the time between COVID-19 onset and the first dose-vaccination spanned around 300 days. HCs and SOTRs were similar in age and gender, and without known morbidities.

**Table I.**
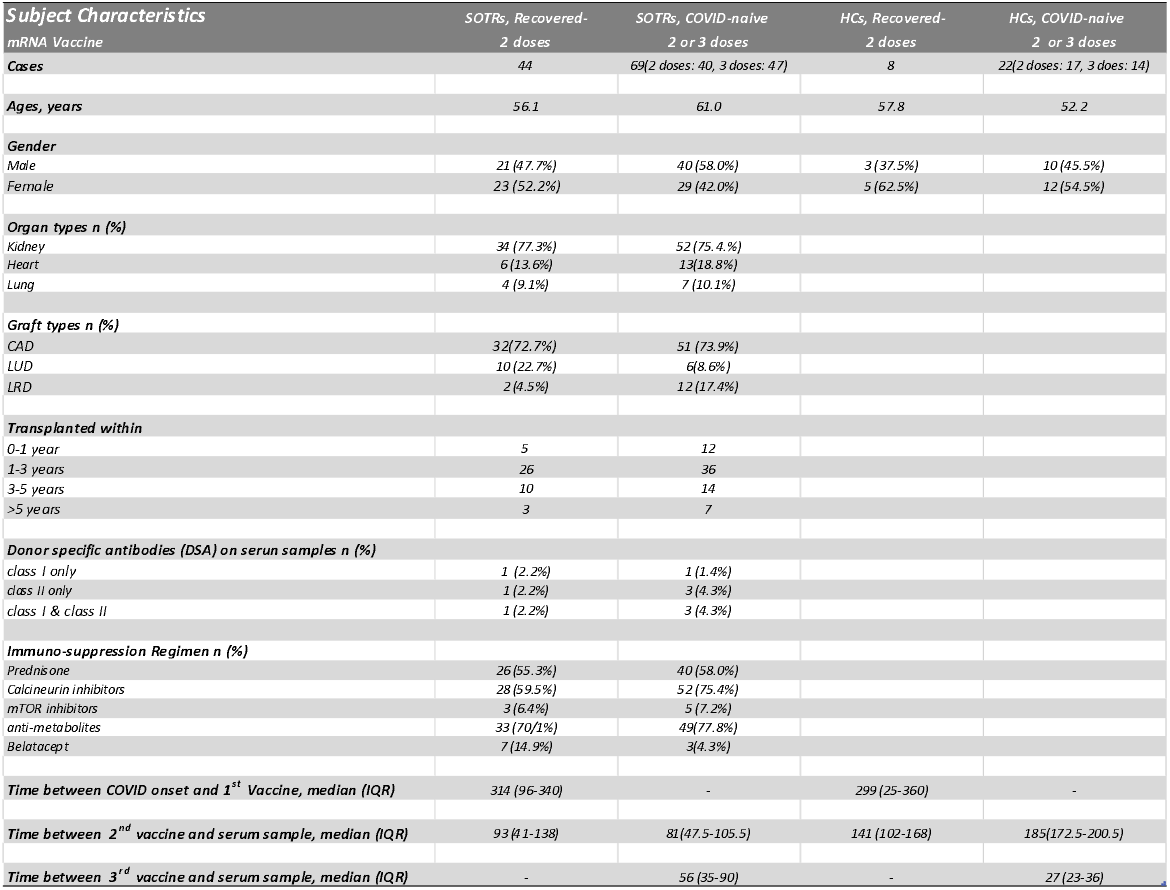
Demographic characteristics of SOTRs and HCs. Note: In the COVID-naïve SOTRs series, sera from 18 subjects were available for both post-2 doses and post-3 dose testing; three patients had two organs transplanted. In the COVID-naïve HCs series, sera from 9 subjects were available for both post-2 doses and post-3 doses testing. Time (days) between the events were presented as median (interquartile range). Abbreviation: CAD, cadaver donor; LUD, living unrelated donor, LRD: living related donor.

### 3.2 Extreme heterogeneity of anti-SARS-CoV-2 immune response in vaccinated SOTRs

The levels of anti-SARS-CoV-2 (nucleocapsid, RBD and S1) IgG antibodies in post-vaccine sera collected from of SOTRs and HCs were analyzed (Figure 1). We found that production of anti-RBD IgG antibodies in COVID-naive SOTRs following vaccination was extremely heterogeneous, compared to their HC counterparts. While HCs were 100% seropositive post-2-dose or post-3-dose vaccination, COVID-naive SOTRs were 27.5% (11/40) seropositive 81 days after 2-dose vaccination, and 55.3% (26/47) seropositive 56 days after 3-dose vaccination. In addition, while the median antibody levels for COVID-naïve HCs was 3,920 (2,668 - 6,568) MFI post-2-dose vaccination and 12,532 (11,583 - 14,956) MFI post-3-dose vaccination, the median antibody levels for COVID-naive SOTRs was 84 (30 - 2170) MFI post-2-dose vaccinations and 1699 (86 - 5806) MFI post- 3-dose vaccination, respectively. The patterns of anti-S1 IgG levels in these patients were similar but were about ½-¼ of that of anti-RBD IgG (data not shown).

**Figure 1:**
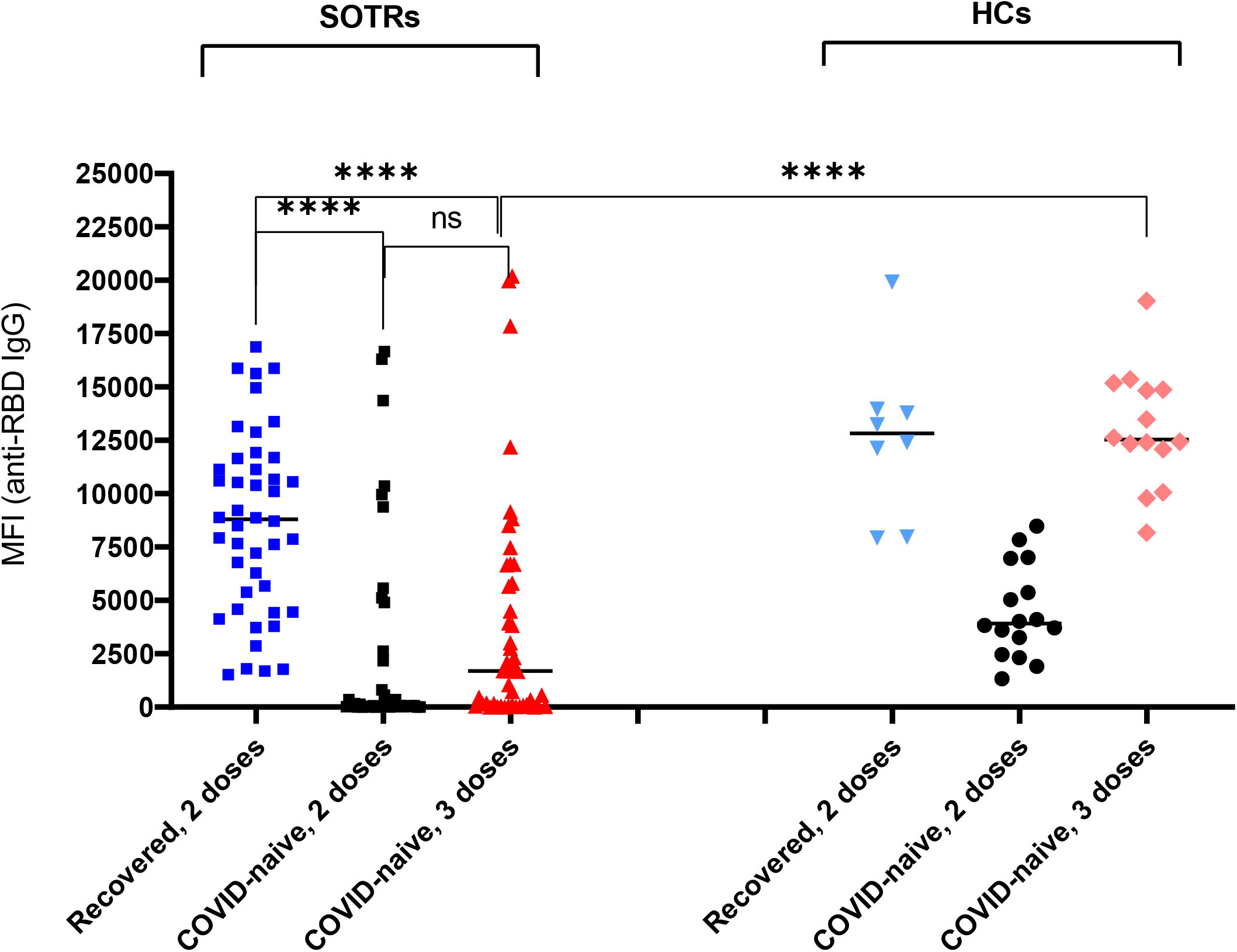
Vaccination induced anti-RBD IgG antibodies in SOTRs and HCs. Sera from recovered SOTR (n = 44) who received 2 doses of vaccines and sera from COVID-naïve (n = 69) who received 2 or 3 doses of vaccines, along with samples from recovered-HCs (n = 8) and COVID-naive HCs (n = 22) were tested for IgG antibodies against SARS-CoV-2 antigens (RBD, S1 and nucleocapsid) by Multiplexed magnetic bead-based assay. Levels of anti-nucleocapsid IgG antibodies were used for confirmation of COVID infection. Positivity (> 700 MFI) of assay, pre-set by manufacturer, is denoted as a horizonal line.

These results reveal the very weak and heterogenous nature of anti-SARS-CoV-2 immune responses in COVID-naïve SOTRs after vaccination. Although there was an increase in the levels and the percentages of anti-RBD IgG seropositivity in COVID-naïve SOTRs after the 3^rd^ dose of vaccination. This was not significantly higher (p = 0.265) than after the 2^nd^ dose; it did, however, remain significantly lower (p < 0.0001) than the response found in HCs.

Analyses of anti-RBD IgG in vaccinated-recovered-SOTRs (n = 44) revealed that these patients mounted a significantly higher immune response against SARS-CoV-2. All 44 patients were anti-RBD seropositive. The median level of IgG antibodies, (8,797 MFI), was significantly higher than that of COVID-naïve who received either 2 dose (p < 0.0001) or 3 dose (p < 0.0001) vaccinations.

### 3.3 Differential neutralization capacities between vaccinated recovered and vaccinated COVID-naïve SOTRs

To determine whether sera from vaccinated SOTRs can neutralize the SARS-COV2 vaccine and VOC, we chose two different approaches. The first was based on the ability of sera to prevent the entry of SARS-CoV-2 pseudotyped lentiviral particles into cells overexpressing ACE2 receptor protein, 293-ACE2 cells. The second was to test the ability of sera to inhibit binding of probed viral Spike proteins to ACE2. For the latter, we opted to use a commercially available multiplexed neutralization system which allows the simultaneous determination of the neutralizing capacities of sera against several SARS-CoV-2 variants. As shown in Figure 2S, we demonstrated that both methods could accurately identify the neutralization capacities against the vaccine and a few (alpha, beta and delta) variant strains on a wide range of clinical diagnostic sera samples. Although multiplexed ACE2 inhibition assay is 10 to 20-fold weaker in sensitivity, it is highly desirable for clinical testing because of its effectiveness as a high-throughput assay.

**Figure 2:**
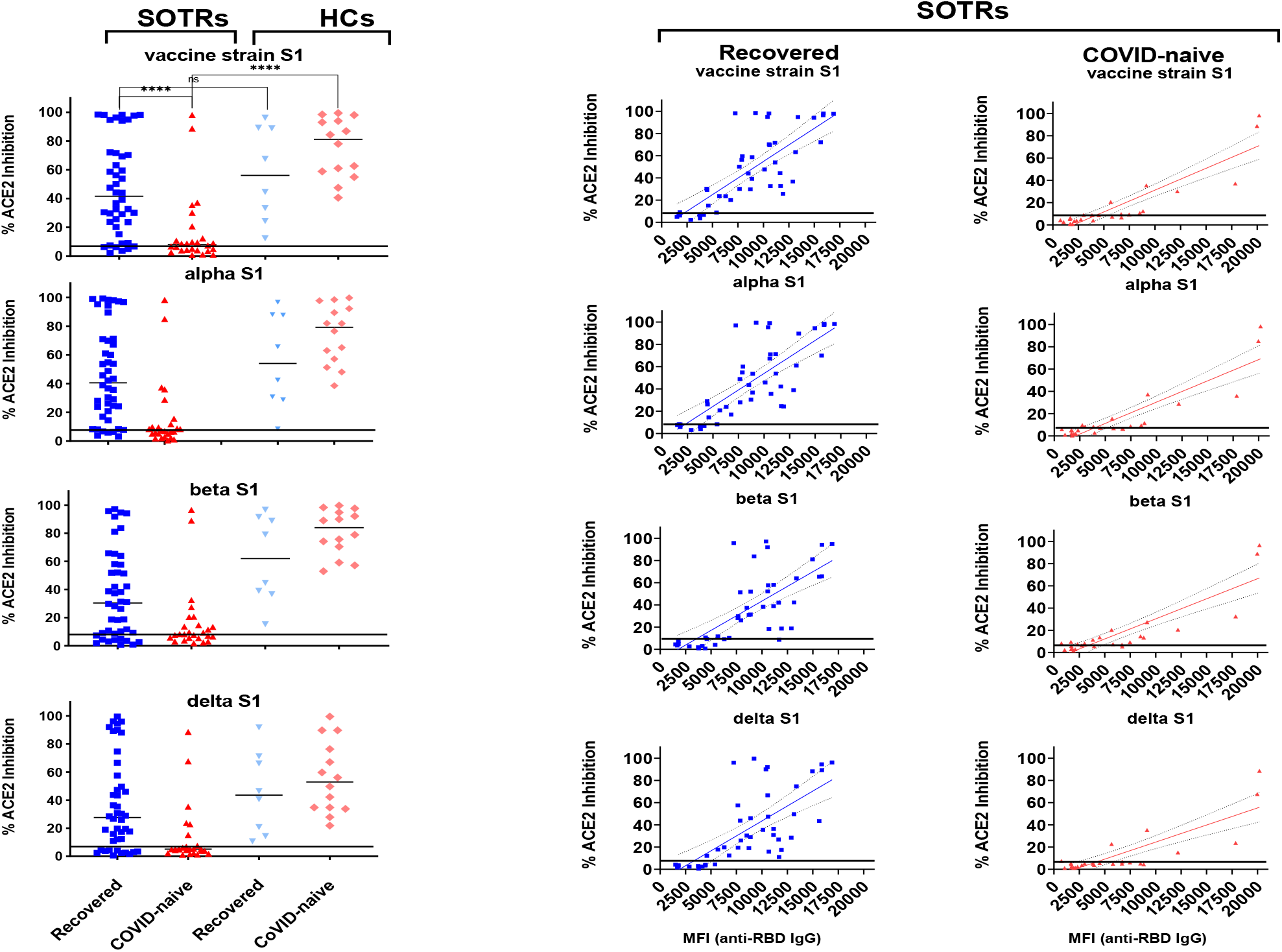
Neutralizing capacities of sera from vaccinated SOTRs or HCs against the vaccine and alpha, beta and delta variants. A: Multiplexed neutralization assays were used to determine the degree of inhibition (%) by sera on binding of indicated viral S1 proteins to ACE2 receptor. A horizonal line (8% inhibition at 1:200-fold dilution) representing the positive neutralizing cutoff was derived from the results of Figure S2. The qualitative analysis (% inhibition) is defined as 100 x (1-sample value /negative control value). B: Liner regression analyses were carried out to determine relationships between the levels of anti-RBD IgG antibodies (X axis) and % ACE2 inhibition (Y axis). In the recovered-SOTRs series, levels of anti-RBD IgG were found strongly correlated with the degree (%) of inhibition on binding of vaccine strain (R^2^ = 0.60; p <0.0001), alpha (R^2^ = 0.59; p <0.0001), beta (R^2^ = 0.51; p <0.0001) and delta (R^2^ = 0.48; p <0.0001) to ACE2. In the COVID-naïve SOTRs series, the correlations between these two events were even stronger, with vaccine strain (R^2^ = 0.80; p < 0.0001), alpha (R^2^ = 0.79; p < 0.0001), beta (R^2^ = 0.75; p < 0.0001), and delta (R^2^ = 0.69; p < 0.0001), respectively.

To determine whether sera from subjects positive for anti-RBD IgG could have neutralizing capacities against the SARS-COV-2 strain and VOC, samples from either fully vaccinated COVID-naïve SOTRs (n = 26) or partially (2-dose) vaccinated recovered-SOTR (n = 44), along with the counterparts from HCs, were measured for their abilities to inhibit viral S1-ACE2 binding. As shown in the panel A of Figure 2, 79.5 % (35/44) of recovered-SOTRs and 61.5 % (16/26) of COVID-naïve SOTRs showed detectable neutralizing activities. If patients with seronegative sera were included, only 34.0 % (16/47) of COVID-naïve SOTRs displayed immune responses against SARS-CoV-2 after 3 doses of vaccination. Median inhibition capacities of sera from recovered-SOTRs, ranged from 27.7 to 41.6% against 4 different viral S1s. These were significantly higher (p < 0.0001) than the results from COVID-naïve SOTRs, which ranged from 5.1 to 8.0%, but were not significantly different from their HC counterparts. Among S1 proteins, the most resistant to neutralization by sera was the delta variant, with a median of 27.7% inhibition for recovered-SOTRs and a median of 5.1% inhibition for COVID-naïve SOTRs. Sera from both HCs groups, however, showed median > 44% cross-strain neutralization capacities.

To further characterize the neutralization patterns occurring in vaccinated recovered-SOTRs and fully vaccinated COVID-naïve SOTRs, we directly compared sera neutralization activities of these two groups of patients against levels of anti-RBD IgG (Figure 2B). We found that neutralization activities against all 4 viral strains were strongly positive-correlated (p < 0.0001) with levels of anti-RBD IgG antibodies in both recovered-SOTRs and COVID-naïve SOTRs. Spearman r ranged from 0.762 to 0.726 for recovered-SOTRs, and from 0.874 to 0.675 for COVID-naïve SOTRs. Our findings suggest that impairment in neutralizing viruses in COVID-naïve SOTRs was due to their inability to produce anti-RBD IgG antibodies, and that boosting anti-viral RBD IgG levels may increase the cross-neutralizing effect against the SARSCoV2 strain and VOC.

### 3.4 Significant escape from neutralization by omicron variant in fully vaccinated COVID-naïve SOTRs

To test if SOTRs positive for anti-RBD IgG antibodies were protected against highly mutated VOC such as omicron, we tested the ability of their sera, as well as of sera from the HCs, to neutralize the omicron variant in pseudotyped neutralization assays. As shown in Figure 3A, we found that fully vaccinated COVID-naïve SOTR displayed extremely heterogenous and low immune responses against omicron. The median level of neutralizing activities (IC50 = 0.3) in this group of individuals was significantly lower than that of fully vaccinated COVID-naïve HCs (IC_50_ = 1582, p <0.0001). Only 4 of 47 of COVID-naïve SOTRs showed neutralizing activity on par with those of HCs. In contrast, vaccinated-recovered SOTR demonstrated higher neutralization activity against omicron. Their neutralizing antibody titers (median IC_50_ = 193) were not significantly different than those of vaccinated recovered-HC (median IC_50_ = 414, p = 0.404) or of fully vaccinated naïve-HCs (IC_50_ = 1582, p = 0.088), but were significantly higher (p = 0.0027) than those of fully vaccinated COVID-naive SOTRs. To better understand the omicron-neutralization patterns observed in sera from these two groups of patients (vaccinated COVID-naïve and vaccinated recovered-SOTRs), we compared their omicron-neutralization with neutralizations against the vaccine strain and the delta variant. We demonstrated that omicron-neutralization was strongly correlated with neutralization against the original strain (Spearman r = 0.870, p < 0.0001) and against delta variant (Spearman r = 0.869, p < 0.0001) in the vaccinated recovered-SOTRs; but were weakly correlated with the neutralization against the vaccine strain (Spearman r = 0.326, p = 0.104) and the delta variant (Spearman r = 0.376, p = 0.058) in the vaccinated COVID-naïve SOTRs (Figure 3B).

**Figure 3:**
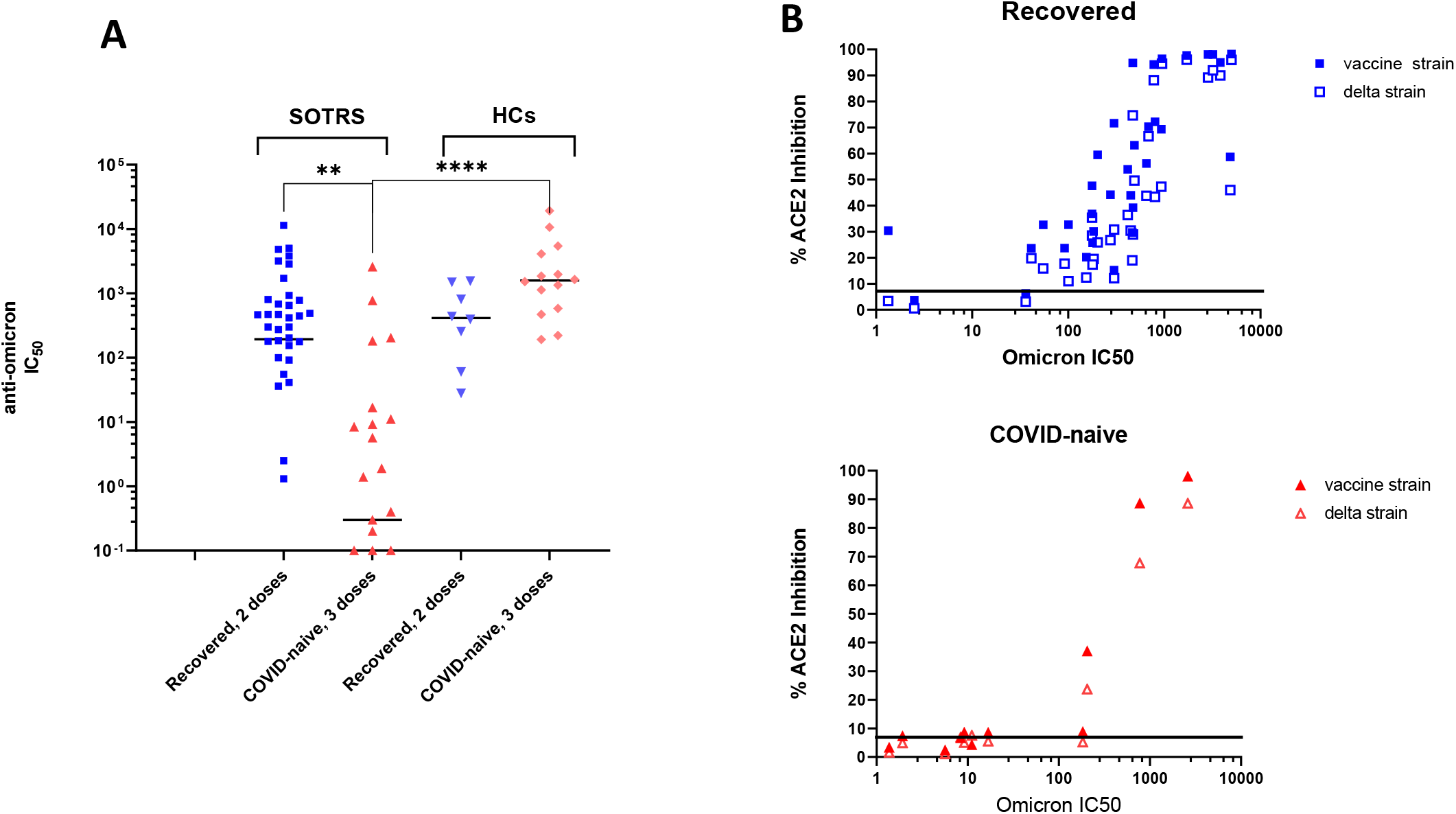
Neutralizing capacities of sera from vaccinated SOTRs and HCs against omicron. A: Sera from recovered-SOTRs or HCs who received 2 doses vaccination and COVID-naïve SOTRs or HCs who received 3 doses of vaccinations were tested for neutralization assays against omicron pseudoviruses. Omicron-neutralization of COVID-naïve SOTRs (median IC_50_ = 0.3) was significantly lower than that of recovered-SOTRs (median IC_50_ =193, p = 0.0027) or COVID-naïve HCs (median IC_50_ = 414, p < 0.0001). B: Correlation tests between omicron-neutralization with vaccine- and delta-neutralization. In the recovered-SOTRs series, omicron-neutralization strongly correlated with both vaccine strain-(Spearman r = 0.870, p<0.0001) and delta strain-neutralization (r = 0.869, p <0.0001). In the COVID-naïve SOTRs series, omicron-neutralization was modestly correlated with both vaccine strain-neutralization (Spearman r = 0.326, p = 0.104) and delta strain-neutralization (Spearman r = 0.376, p = 0.058).

## Discussions

In this study, we described humoral responses to COVID-19 mRNA vaccination from two groups of SOTRs. One group consisted of patients previously infected with SARS-CoV-2 who received two doses of vaccinations, while the other consisted of patients who did not contract the virus but were fully vaccinated. Our results suggest that previous COVID infection not only triggers the induction of higher titers of neutralizing antibodies in SOTRs against both the original strain and lesser-mutated delta variant but raises the breadth of overall humoral immunity reflected in cross-reactivities, allowing efficient neutralization against a highly mutated variant such as omicron. A vast majority of fully vaccinated COVID-naïve SOTRs, on the other hand, failed to do so. Our study provides some important insights into the interplay between naturally acquired and vaccine induced antiviral immune responses in SOTRs.

Compared to HCs, humoral responses against COVID-19 vaccine in the COVID-naïve group were low and very heterogeneous. While a booster (3^rd^ dose) greatly increased the levels of anti-RBD IgG antibodies and cross-variant neutralizing capacities in all of HCs, only a minority (16/47) of SOTRs responded to the booster. A majority (> 65%) of COVID-naïve SOTRs, however, produced either no or very limited levels of anti-RBD IgG antibodies and hence no or sporadic levels of neutralizing capacities against past-dominant (alpha, delta) or current-dominant (omicron) strains. It is not surprising, but alarming, that only 4 at of a total of 47 COVID-naïve SOTRs cohort produced levels of omicron-neutralizing antibodies comparable to those from HCs. Our finding appears to be in an agreement with a recent study by Benning et al^17^, who demonstrated that vaccine-induced cross-neutralization against omicron was detectable in 43% (15/35) anti-S1 IgG-seropositive kidney transplant patients. Omicron variants have been shown to be well-equipped to escape from neutralization by many therapeutic monoclonal antibodies, yet retain a high affinity for the ACE2 receptor^18,19^.

Our study showed that the majority of recovered-SOTRs maintained a robust anti-viral response, with respect to levels of anti-RBD antibodies and cross-variant neutralizing capacities, to levels almost on par with those of HCs. Given that both SOTR groups received similar immunosuppressant regimens and patient care, it is striking to note that many SOTRs who contracted SARS-CoV-2 during the very early days of pandemic, regained strong anti-viral immune response upon vaccination. We speculate that naturally acquired anti-SARS-CoV-2 immune responses via infection, even a mild one, were long-lasting and more stimulatory than the vaccine induced immunities, as demonstrated by others^20^. The effectiveness of T cell-dependent antibody responses is optimized by preferentially steering B cells reactive against high affinity or abundant epitopes toward plasma cell differentiation^21^. Processing and presentation of multiple HLA-bound viral epitopes by macrophages and dendritic cells may contribute to intra- and inter-molecular spreading of cryptic epitopes. This may explain the broad spectrum of anti-variant antibodies in COVID–19 recovered patients ^22,23^.

As increasing numbers of individuals in the SOTR population are infected, tailor-made health plans for each patient may be needed. Our study provides a glimpse into the difference of vaccine responses in SOTRs, with versus without a history of COVID-19 infection. These data indicate that the 3-dose MRNA vaccine is likely insufficient to provide protection against severe COVID in many SOTRs, underscoring the need to maintain preventative measures after vaccination involving social distancing or shielding. Additional studies of humoral responses to extended vaccines also need to be done.

## Limitation of the study

The study is from a single-center and has a retrospective design with a small cohort. Due to the laboratory’s setting, we were not able to carry out the live-virus neutralization assays to confirm results obtained from pseudotyped neutralization or ACE2 inhibition assays.

## Data Availability

All data produced in the present study are available upon reasonable request to the authors

## Abbreviations

SOTR: solid organ transplant recipient
HC: healthy control
SARS-CoV-2: Severe Acute Respiratory Syndrome Coronavirus 2
COVID-19: coronavirus disease 2019
VOC: variant of concern
RT-PCR: Real-Time polymerase chain reaction
RBD: receptor binding domain
MFI: mean fluorescence intensity
IC_50_: the half maximal inhibitory concentration
IQR: interquartile range

## ACKNOWLEDGENTS

We thank every member of laboratory of Immunogenetics for encouragement and collection of samples. CC specially wants to thank his wife and children for their love and support during the most critical stage of his life. CC is also very grateful to the doctors and nurses at White Plains Hospital, Burke Rehabilitation Hospital, Dr. Min-Tsung Hsu and Dr. Mike Liaw for their dedication and excellent patient care.

The work was supported by intramural research Programs of Department of Pathology and Cell Biology, Columbia University.

## Figure Legends

**Figure S1:**
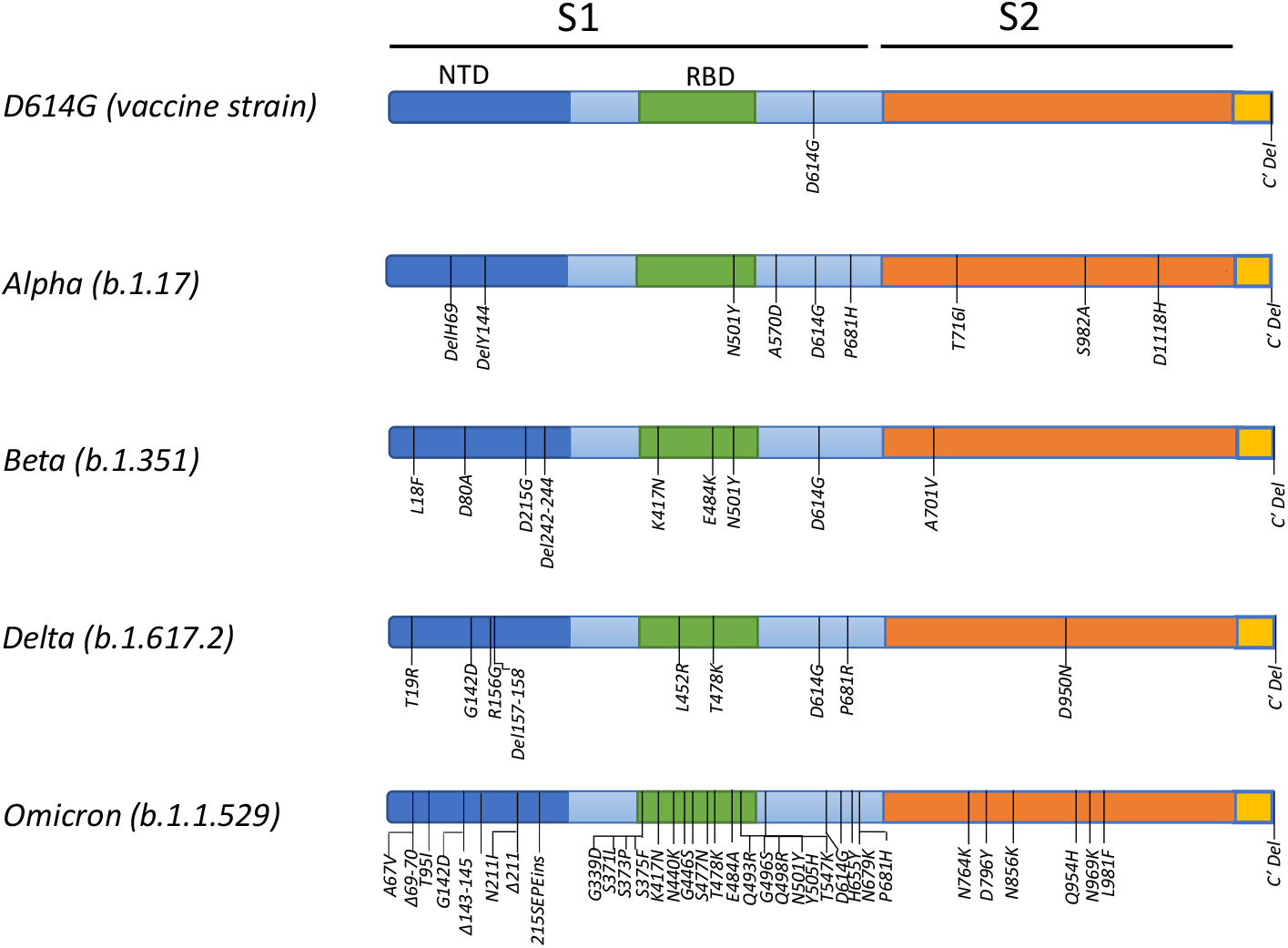
Diagram illustrating the spike proteins of the vaccine strain, alpha, beta, delta and omicron variants. Mutations present in each of the spike protein variants are labeled. Since all known variants carrying the D614G mutation in the non-receptor binding domain region of S1 subunit, the original D614G strain was used as the representative of the vaccine strain.

**Figure S2:**
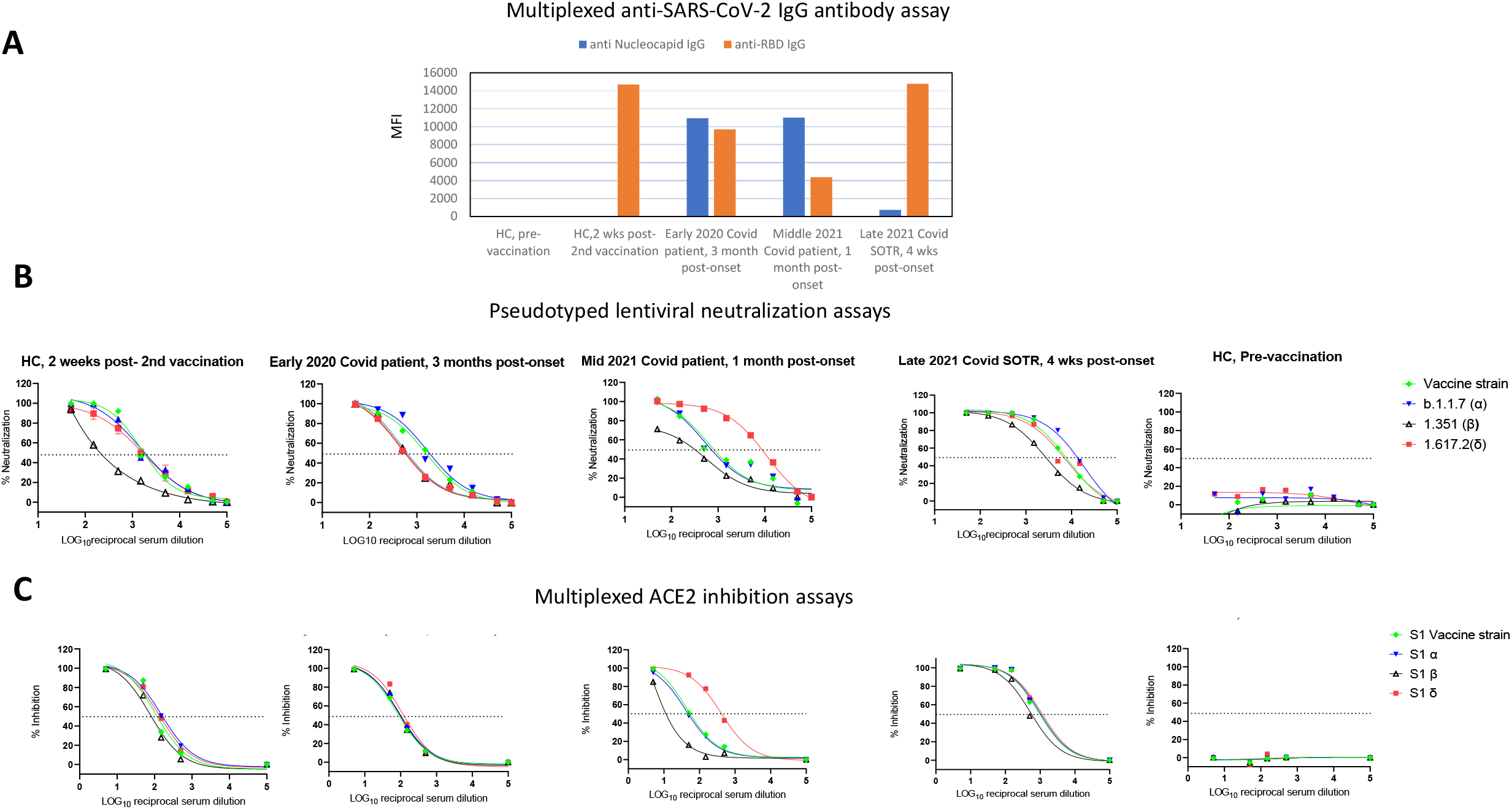
Detection of neutralizing activities in vaccinated or SARS-CoV-2 infected individuals. **A)** A panel of diagnostic serum samples (two samples, pre-vaccinated and post 2 doses vaccinated, from a healthy control; one from an early 2020 COVID patient; one from a mid-2021 COVID patient and one from a late 2021 breakthrough-COVID SOTR) were measured for anti-SARS-CoV-2 IgG, pseudotyped neutralization assays and multiplexed ACE2 inhibition assays. **B**) SARS-CoV-2 pseudotyped neutralization assays were performed with a serially (50-50,000-fold) diluted serum. The sample from late 2021 breakthrough-COVID SOTR had the highest overall IC_50_ values, ranging from highest (IC_50_ = 12,312) against alpha S1 to lowest (IC_50_ = 2,617) against beta S1, whereas the sample from mid-2021 COVID subject showed a skewed anti-viral immune response, with strongest activities (IC_50_ = 7861) against the delta variant and weakest activities (IC_50_ = 253) against the beta variant. This typical immune response has been described by others^14^. **C**) ACE2 inhibition assays performed with a serially (5-5,000) diluted sera used in the Panel B. Average median IC_50_ against all 4 viral S1 proteins for all samples was 114.5 (79.9 -457). Similar to Panel B, ACE2 inhibition assays demonstrated that serum from the breakthrough-SOTR had the highest antiviral immune activities and serum from mid-2021 COVID subject had a skewed anti-viral immune response. Based on this result, we diluted majorities of sera 200-fold for this assay. No pre-existing anti-SARS-CoV-2 IgG, or neutralizing activities were found in serum from the pre-vaccinated individual.

## DATA AVAILABILITY STATEMENT

The data that support the finding of this study are available from the corresponding author upon reasonable request.

